# Modeling effectiveness of testing strategies to prevent COVID-19 in nursing homes —United States, 2020

**DOI:** 10.1101/2020.12.18.20248255

**Authors:** Isaac See, Prabasaj Paul, Rachel B. Slayton, Molly K. Steele, Matthew J. Stuckey, Lindsey Duca, Arjun Srinivasan, Nimalie Stone, John A. Jernigan, Sujan C. Reddy

## Abstract

**Background:** SARS-CoV-2 outbreaks in nursing homes can be large with high case fatality. Identifying asymptomatic individuals early through serial testing is recommended to control COVID-19 in nursing homes, both in response to an outbreak (“outbreak testing” of residents and healthcare personnel) and in facilities without outbreaks (“non-outbreak testing” of healthcare personnel). The effectiveness of outbreak testing and isolation with or without non-outbreak testing was evaluated.

**Methods:** Using published SARS-CoV-2 transmission parameters, the fraction of SARS-CoV-2 transmissions prevented through serial testing (weekly, every three days, or daily) and isolation of asymptomatic persons compared to symptom-based testing and isolation was evaluated through mathematical modeling using a Reed-Frost model to estimate the percentage of cases prevented (i.e., “effectiveness”) through either outbreak testing alone or outbreak plus non-outbreak testing. The potential effect of simultaneous decreases (by 10%) in the effectiveness of isolating infected individuals when instituting testing strategies was also evaluated.

**Results:** Modeling suggests that outbreak testing could prevent 54% (weekly testing with 48-hour test turnaround) to 92% (daily testing with immediate results and 50% relative sensitivity) of SARS-CoV-2 infections. Adding non-outbreak testing could prevent up to an additional 8% of SARS-CoV-2 infections (depending on test frequency and turnaround time). However, added benefits of non-outbreak testing were mostly negated if accompanied by decreases in infection control practice.

**Conclusions:** When combined with high-quality infection control practices, outbreak testing could be an effective approach to preventing COVID-19 in nursing homes, particularly if optimized through increased test frequency and use of tests with rapid turnaround.

**Summary:** Mathematical modeling evaluated the effectiveness of serially testing asymptomatic persons in a nursing home in response to a SARS-CoV-2 outbreak with or without serial testing of asymptomatic staff in the absence of known SARS-CoV-2 infections.

## Introduction

Coronavirus disease 2019 (COVID-19) has posed a significant public health challenge for nursing homes in the United States [1,2]. To prevent introduction and transmission of SARS-CoV-2, nursing homes have been recommended to implement precautions including monitoring symptoms of residents and healthcare personnel (HCP), testing symptomatic persons promptly, and instituting infection control practices including, but not limited to, isolating individuals with COVID-19 [3]. In addition, because asymptomatic and pre-symptomatic infected individuals can transmit SARS-CoV-2, identifying such infected individuals early through serial testing is also recommended to control COVID-19 in nursing homes both as part of an outbreak response as well as in facilities not experiencing outbreaks [4–6].When responding to an outbreak, CDC currently recommends testing all residents and HCP every 3–7 days until no new cases are identified. In addition, in facilities not experiencing an outbreak, serial testing of all HCP is currently recommended, at intervals dependent on the level of county transmission [6].

Infection prevention and control strategies in nursing homes might be hampered by shortages of testing supplies [7] and the time-consuming effort needed to implement widespread testing strategies. The impact of various testing strategies for asymptomatic residents and HCP has not been well characterized; understanding the relative benefit of testing strategies may help prioritize resources. The objective of this manuscript is to provide estimates of the effectiveness (i.e., percentage of COVID-19 cases prevented) of either outbreak testing or combining outbreak and non-outbreak testing for preventing COVID-19 in nursing homes under different scenarios of test frequency and performance. We also estimate the “efficiency” (number of tests needed to prevent a case) of both outbreak and non-outbreak testing strategies.

## Methods

### Definitions

“Outbreak testing” was defined as serial testing (e.g., daily, every three days, weekly) of all residents and HCP immediately following recognition of an initial COVID-19 case. “Non-outbreak testing” was defined as serial testing of HCP in the absence of a known COVID-19 case. Mathematical modeling was used to evaluate two paradigms for using testing to help control COVID-19 in nursing homes: (1) outbreak testing that begins immediately following recognition of an initial case, and (2) non-outbreak testing in facilities when outbreaks are not occurring combined with outbreak testing once cases are identified. Testing frequency, turnaround time, and sensitivity were varied to broadly reflect testing capabilities as of November 2020. The primary quantities of interest were the “effectiveness” of a testing strategy, defined as the percent of cases prevented; and the “efficiency” of a testing strategy, defined as the number of cases prevented divided by the number of tests used.

Evaluating these strategies for a “typical” nursing home required estimating three quantities through modeling: (1) the expected number of SARS-CoV-2 infections occurring in an outbreak without testing of asymptomatic persons, i.e., infection control is guided only by testing and isolating symptomatic residents and HCP. This quantity is considered the “baseline” outbreak size because, conceptually, our evaluation is for paradigms of testing asymptomatic persons. (2) the expected number of SARS-CoV-2 infections occurring per outbreak when outbreak testing is conducted. The difference between (2) and (1) is the number of cases prevented by outbreak testing.(3) the expected percentage of SARS-CoV-2 outbreaks prevented by non-outbreak testing.

Parameters used for the model and estimates were set as follows: during the early months of the pandemic the mean number of residents from Centers for Medicare & Medicaid Services (CMS)-certified nursing homes was 86 [8]. The HCP per resident ratio was set at 1.5:1, yielding 129 HCP at the facility based on the following rationale: analysis of the payroll-based journal data from the Centers for Medicare and Medicaid Services (PBJ) suggest that on average 1.2 full-time equivalent HCP per resident are employed by each nursing home [9]. However, this likely represents a minimum estimate because some HCP work part-time. In some areas where facility-wide testing of and residents has been conducted, the HCP per resident ratio has been as high as 2:1 (CDC, unpublished data). The ratio of 1.5 HCP per resident was chosen as closer to the midpoint of these estimates. Although these HCP estimates HCP may include some who do not have direct resident contact, such HCP could still contribute to SARS-CoV-2 transmission (e.g., HCP-to-HCP). Estimates of infectivity from asymptomatic or pre-symptomatic persons [10] varied over the course of illness and were used to calculate the expected reduction of transmission occurring from detecting these people through testing and isolating them (supplemental methods). All scenarios model introductions of SARS-CoV-2 into the facility through HCP only, e.g., no visitors are permitted. A summary of model inputs can be found in Table 1.

**Table 1:** Model inputs

| Variable | Value | Source/reference |
| --- | --- | --- |
| Number of residents | 86 | Estimated as occupied beds from Nursing home compare [8] |
| Number of healthcare personnel | 129 | 1.5 times the number of residents |
| Total population size | 215 | Number of residents+healthcare personnel |
| $R_0$ | 1.366689 | Mean $R_0$ estimated from outbreak sizes among Colorado nursing homes (Supplemental methods) |
| Proportion of transmission from symptomatic infections occurring during presymptomatic phase | 0.5 | CDC Pandemic Planning Scenarios [10] |
| Proportion of infections that are asymptomatic | 0.4 | CDC Pandemic Planning Scenarios [10] |
| Infectivity of asymptomatic versus symptomatic infections | 0.75 | CDC Pandemic Planning Scenarios [10] |

### Calculation of estimates

To estimate the expected size of an outbreak at baseline, a Reed-Frost model, in which the susceptible individuals in a population of size *N* have probability 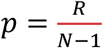 of becoming infected at each stage of transmission [11], was used to model the transmission of SARS-CoV-2 during a nursing home outbreak. The Reed-Frost model was chosen because it is well-suited for capturing stochasticity of transmissions in finite-sized congregate settings. The total population size is the number of residents plus the number of HCP. We estimated the reproductive number R_0_ for outbreaks under the following conditions: (1) the only testing occurring was of symptomatic persons, (2) isolation of symptomatic persons occurred, and (3) recommendations for other established infection control practices (e.g., isolating patients, universal facemasks and cloth face coverings) were in place. This estimate of R_0_, based on data for nursing home outbreaks from Colorado during the early period of the pandemic (supplemental methods), is referred to as the “unmitigated” R_0_ and used to generate the baseline outbreak size.

To determine outbreak size when using outbreak testing, modeling based on viral dynamics of SARS-CoV-2 was used to estimate the reduction in transmission μ expected from the testing strategy (supplemental methods). The mitigated reproductive number R_0_’ was calculated by multiplying the unmitigated R_0_ by the reduction in transmission μ expected from the testing strategy and used to estimate outbreak size using outbreak testing.

For non-outbreak testing, the proportion of outbreaks prevented was set to equal μ. This corresponds to an assumption that an outbreak (at baseline) will typically be introduced by a single person so that the proportion of transmissions prevented equals the proportion of outbreaks averted and represents a best-case scenario for non-outbreak testing regardless of the level of community transmission. As outlined in the supplemental methods, this was combined with estimates for outbreak testing to determine the effectiveness and efficiency of using both outbreak and non-outbreak testing.

The “baseline” test sensitivity was modeled to vary during the course of illness with test sensitivity proportional to infectiousness early in the course of illness (supplemental methods). Peak sensitivity was set at 95% reflecting best understanding of reported reverse-transcriptase polymerase chain reaction (RT-PCR) test characteristics [12]. To mirror realistic and recommended tests available, outbreak and non-outbreak testing strategies were primarily evaluated with weekly and every-three-days testing intervals with either a 24-hour or 48-hour turnaround time and sensitivity corresponding to RT-PCR; or an estimated sensitivity of 85% (compared to RT-PCR tests), with immediate turnaround time, i.e., point-of-care test (supplemental methods). The lower sensitivity of 85% was chosen to match the lower end of the range of sensitivity reported for currently available point-of-care tests [13]. We also evaluated a hypothetical outbreak and non-outbreak testing strategy in which a point-of-care test with 50% sensitivity is used for daily testing. These testing scenarios with lower sensitivity were evaluated because some investigators have advocated for the development and adoption of rapid tests with lower sensitivity [14]. We also evaluated weekly and every-three-days testing with a lower sensitivity (50%) point-of-care platform because of reports that existing point-of-care tests may have lower sensitivity for detecting asymptomatic individuals [15].

### Sensitivity analyses and other variations

The above analyses evaluated the impact of adding non-outbreak testing to outbreak testing with a 10% probability of SARS-CoV-2 introduction to a facility during the week that testing occurs, which corresponded to the overall national picture during July-September 2020 (supplemental methods). As a sensitivity analysis, this probability was increased to evaluate the potential impact of non-outbreak testing in areas with higher levels of community transmission.

We considered that implementing a more comprehensive testing strategy require additional resources (e.g., staff time, protective equipment). We postulated that these increases might have a deleterious effect on a facility’s ability to control transmission through adherence to infection control protocols. To estimate this effect, we calculated the impact of a relatively small (i.e., 10%) decrease (from a default of 100%) in the effectiveness of preventing transmission by isolating infected persons when adding non-outbreak testing to outbreak testing.

Additional sensitivity analyses used adaptations of three alternate infectivity profiles from the literature to calculate the above quantities (supplemental methods; supplemental table S1).

### Administrative information

Modeling calculations were conducted with the statistical software R (version 4.0.2, the R Foundation for Statistical Computing). Additional details about modeling can be found in a supplemental section. Code for the models can be found on the CDC Epidemic Prediction Initiative GitHub site (https://github.com/cdcepi/Nursing-home-SARS-CoV-2-testing-model/).

This activity was reviewed by CDC and was conducted consistent with applicable federal law and CDC policy (see e.g., 45 C.F.R. part 46, 21 C.F.R. part 56; 42 U.S.C. §241(d); 5 U.S.C §552a; 44 U.S.C. §351 et seq.).

## Results

### Effectiveness and efficiency of outbreak and non-outbreak testing

At all currently recommended testing intervals with acceptable turnaround times (i.e., 48 hours or less), outbreak testing alone is estimated to prevent 54% (weekly testing with 48-hour turnaround time) to 90% of cases (point-of-care testing every three days) associated with a COVID-19 outbreak (Table 2). Adding weekly or every-three-days non-outbreak testing to outbreak testing prevented up to 8% of additional cases. Combining every-three-days outbreak and non-outbreak testing with a point-of-care test prevented 95% of cases. In addition, at a given testing frequency and turnaround time, increasing either the frequency of the testing or decreasing the turnaround time for outbreak testing was estimated to prevent more infections than adding non-outbreak testing with the same frequency and turnaround time. In an exploratory analysis, use of a daily POC test with 50% sensitivity was estimated to prevent 92% of cases using outbreak testing alone and 97% of cases using both outbreak and non-outbreak testing.

**Table 2:**
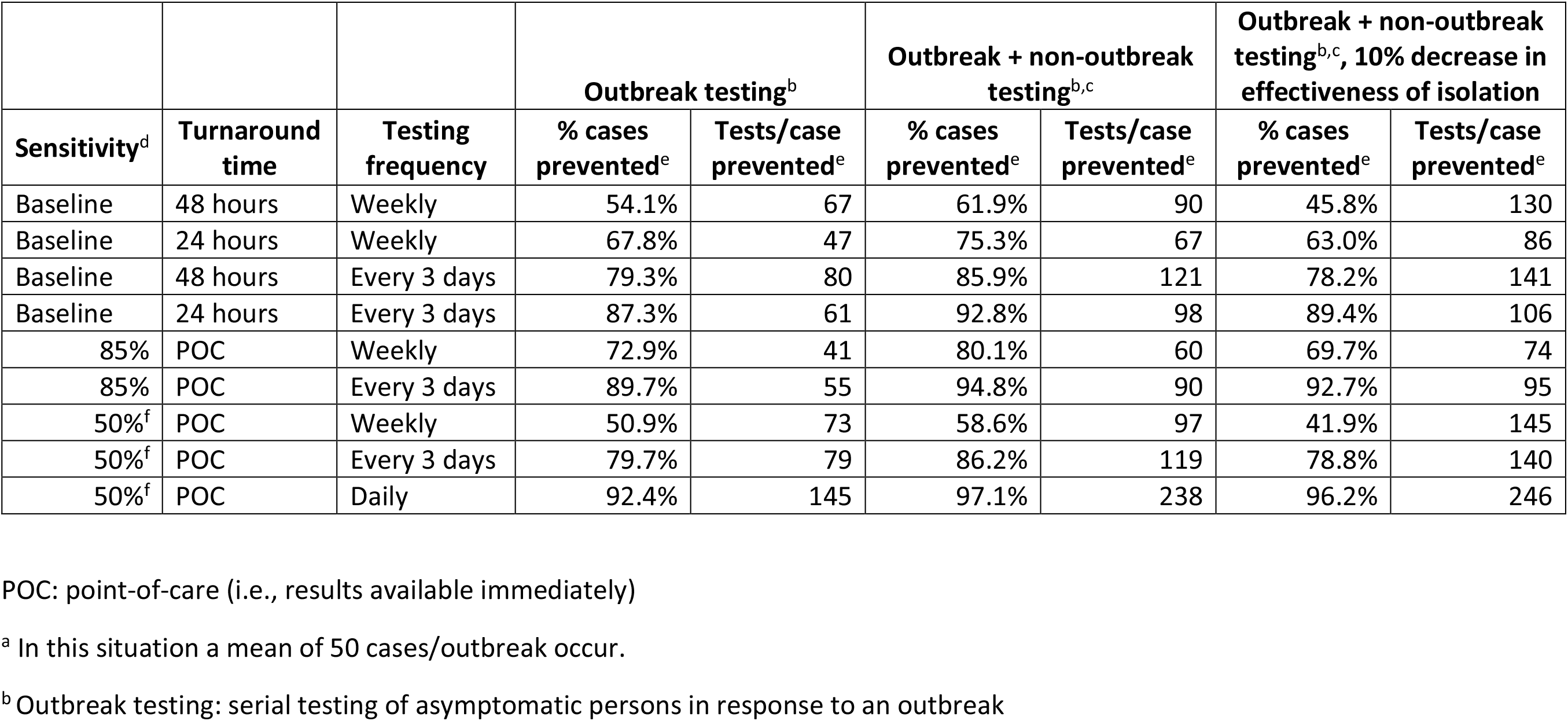
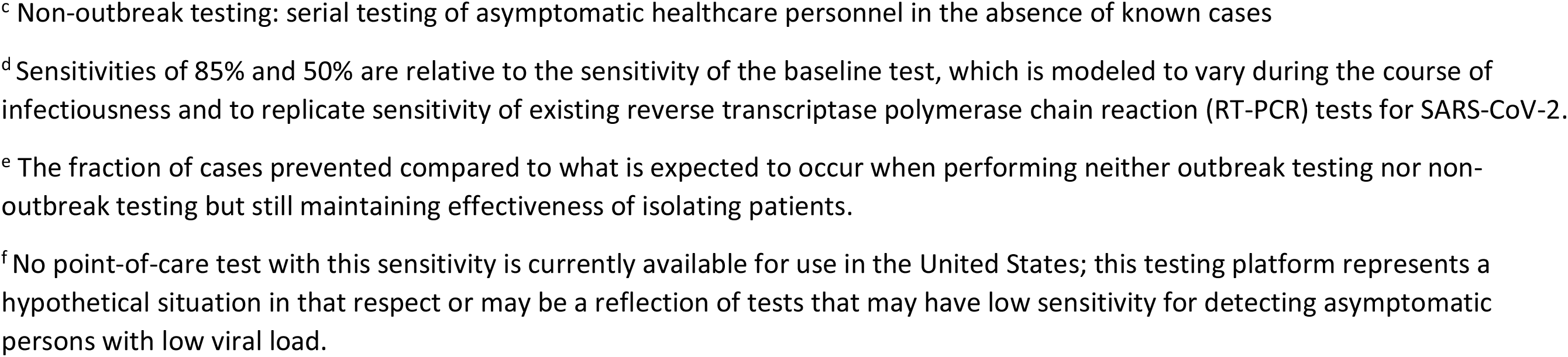
Percent SARS-CoV-2 infections prevented and number of tests needed per case prevented when serially testing asymptomatic nursing home residents and staff in response to outbreaks (“outbreak testing”), combining this testing strategy with serial testing of asymptomatic staff in the absence of known cases (“outbreak+non-outbreak testing”), and conducting both outbreak and non-outbreak testing with a concomitant 10% decrease in the effectiveness of halting SARS-CoV-2 transmission through isolating identified cases. Cases occurring using these strategies are compared to the number occurring with use of only symptom-based testing and isolation^a^.

|  |  |  | Outbreak testing<sup>b</sup> |  | Outbreak + non-outbreak testing<sup>b,c</sup> |  | Outbreak + non-outbreak testing<sup>b,c</sup>, 10% decrease in effectiveness of isolation |  |
| --- | --- | --- | --- | --- | --- | --- | --- | --- |
| Sensitivity<sup>d</sup> | Turnaround time | Testing frequency | % cases prevented<sup>e</sup> | Tests/case prevented<sup>e</sup> | % cases prevented<sup>e</sup> | Tests/case prevented<sup>e</sup> | % cases prevented<sup>e</sup> | Tests/case prevented<sup>e</sup> |
| Baseline | 48 hours | Weekly | 54.1% | 67 | 61.9% | 90 | 45.8% | 130 |
| Baseline | 24 hours | Weekly | 67.8% | 47 | 75.3% | 67 | 63.0% | 86 |
| Baseline | 48 hours | Every 3 days | 79.3% | 80 | 85.9% | 121 | 78.2% | 141 |
| Baseline | 24 hours | Every 3 days | 87.3% | 61 | 92.8% | 98 | 89.4% | 106 |
| 85% | POC | Weekly | 72.9% | 41 | 80.1% | 60 | 69.7% | 74 |
| 85% | POC | Every 3 days | 89.7% | 55 | 94.8% | 90 | 92.7% | 95 |
| 50%<sup>f</sup> | POC | Weekly | 50.9% | 73 | 58.6% | 97 | 41.9% | 145 |
| 50%<sup>f</sup> | POC | Every 3 days | 79.7% | 79 | 86.2% | 119 | 78.8% | 140 |
| 50%<sup>f</sup> | POC | Daily | 92.4% | 145 | 97.1% | 238 | 96.2% | 246 |

POC: point-of-care (i.e., results available immediately)
<sup>a</sup> In this situation a mean of 50 cases/outbreak occur.
<sup>b</sup> Outbreak testing: serial testing of asymptomatic persons in response to an outbreak
<sup>c</sup> Non-outbreak testing: serial testing of asymptomatic healthcare personnel in the absence of known cases
<sup>d</sup> Sensitivities of 85% and 50% are relative to the sensitivity of the baseline test, which is modeled to vary during the course of infectiousness and to replicate sensitivity of existing reverse transcriptase polymerase chain reaction (RT-PCR) tests for SARS-CoV-2.
<sup>e</sup> The fraction of cases prevented compared to what is expected to occur when performing neither outbreak testing nor non-outbreak testing but still maintaining effectiveness of isolating patients.
<sup>f</sup> No point-of-care test with this sensitivity is currently available for use in the United States; this testing platform represents a hypothetical situation in that respect or may be a reflection of tests that may have low sensitivity for detecting asymptomatic persons with low viral load.

Combining outbreak and non-outbreak testing in the setting of a 10% decrease in the effectiveness of isolating infected persons prevented fewer cases than outbreak testing alone with complete effectiveness of isolation for all testing frequencies, turnaround, and test sensitivity scenarios evaluated except when testing every three days with 24-hour turnaround time or POC testing with 85% sensitivity. In these scenarios, adding non-outbreak testing but decreasing effectiveness of isolating infected persons was estimated to prevent an additional 2-3% of cases compared to outbreak testing alone with complete effectiveness of isolation.

The number of tests needed to prevent a case through outbreak testing alone (range: 41-145 tests/case prevented) was lower than through combining outbreak and non-outbreak testing (range: 60-238).

### Varying likelihood of SARS-CoV-2 introduction from the community

When conducting outbreak and non-outbreak testing every three days with a point-of-care test with 85% sensitivity, as the likelihood of SARS-CoV-2 introductions per week increased, the percent of cases prevented was estimated to remain unchanged at 95% (Figure). However, as the likelihood of SARS-CoV-2 introductions increased, the number of tests needed to prevent a case was estimated to decrease from 90 tests/case prevented at a 10% likelihood of introductions to 9 tests/case prevented at a 100% likelihood of introductions when combining outbreak and non-outbreak testing (Figure).

**Figure:**
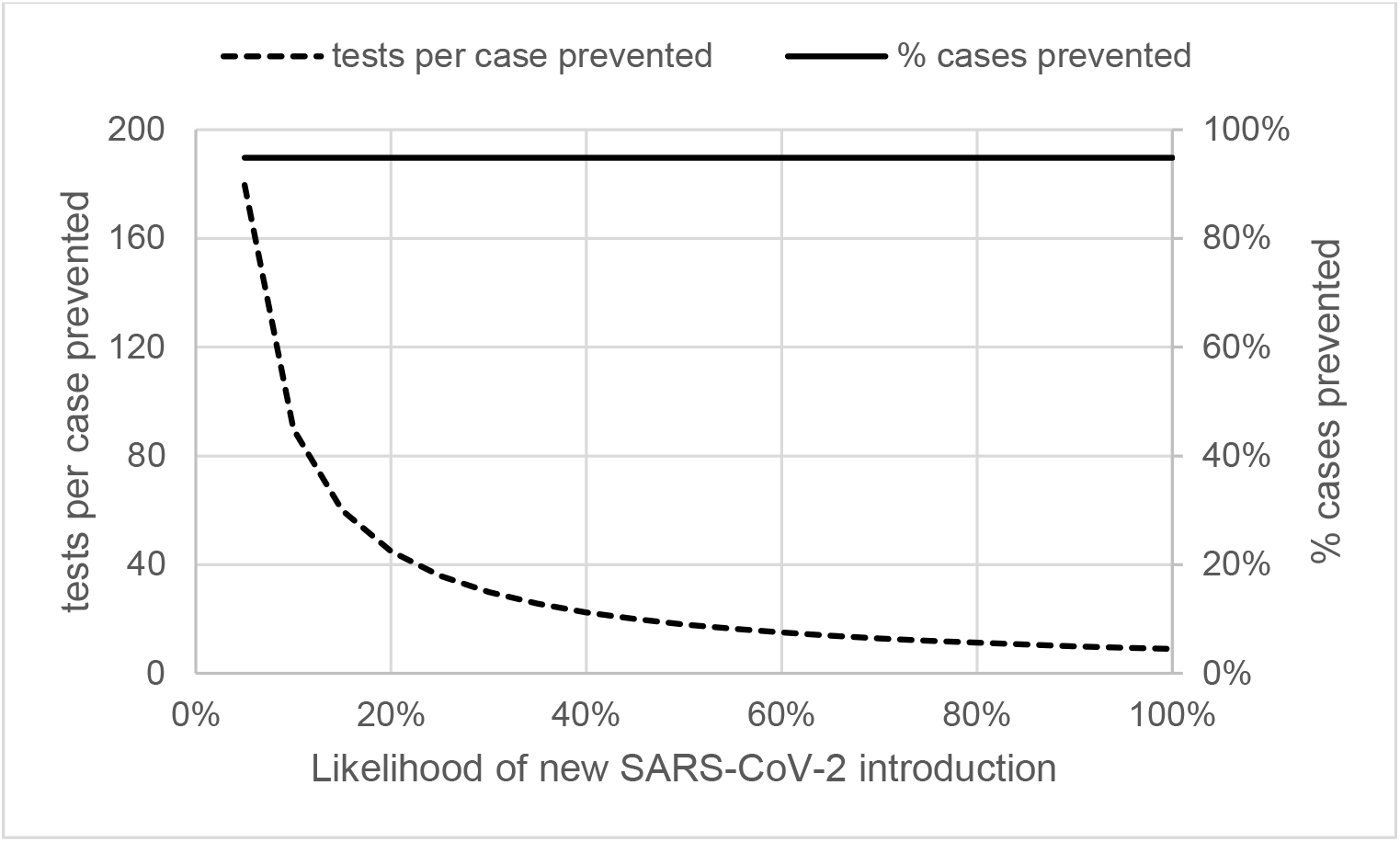
Evaluation of how the performance of testing strategies for nursing homes changes as the likelihood of a new SARS-CoV-2 introduction during the week of testing varies. The solid line depicts the percentage of cases prevented when combining outbreak and non-outbreak testing. The dotted line shows tests per case prevented when combining outbreak and non-outbreak testing. Both lines show results for outbreak and non-outbreak testing conducted every 3 days with immediate turnaround (i.e., point-of-care test) and 85% test sensitivity (compared to a reverse-transcriptase polymerase chain reaction [RT-PCR] test).

### Sensitivity analyses

As a sensitivity analysis when using alternative infectivity profiles (Table S1), the magnitude of benefit of outbreak testing varied (Tables S2-S4). For example, testing weekly with 48-hour turnaround prevented 31–74% of infections depending on the infectivity profile. However, in all of these sensitivity analyses adding non-outbreak testing added a relatively small percentage benefit to outbreak testing alone, not exceeding 8% for any test turnaround/frequency/sensitivity regardless of infectivity profile. In addition, for all but one infectivity profile (Table S4), for the majority of testing scenarios modeled outbreak testing alone was superior to combining outbreak and non-outbreak testing if combined testing was associated with decreases in effectiveness of isolating patients.

## Discussion

Preventing COVID-19 in nursing homes is a high priority for the pandemic response [16]. In our analysis, serially testing asymptomatic residents and HCP in response to an outbreak to guide infection control response prevented the majority of SARS-CoV-2 infections in a hypothetical nursing home with 86 residents and 129 staff. In addition, optimizing and prioritizing outbreak testing to facilitate increased test frequency with rapid turnaround times while maintaining high-quality infection control practice is estimated to be an efficient strategy while also nearly maximizing number of infections prevented.

Ensuring robust symptom identification with prompt testing in response to a potential new SARS-CoV-2 infection, and subsequently eliminating transmission from persons with COVID-19 through isolation, is essential for responses using outbreak testing to be effective. Nursing homes with outbreaks frequently face staff shortages [17], which may limit capacity to perform necessary patient care activities, infection prevention activities, or increased testing. If redirecting resources from recommended infection control practices to testing leads to small reductions in the effectiveness of infection control practices, then the combination of outbreak and non-outbreak testing could be less effective than just performing outbreak testing with high-quality infection control. These considerations are all the more important because real-life conditions in a nursing home might not correspond to ideal implementation. For example, isolation of patients can be difficult if there are no empty rooms; and isolated patients still need care and therefore could still pose some risk of transmission to staff.

These findings also support current recommendations from CDC and CMS [6,18] to prioritize testing of symptomatic residents and HCP first, followed by testing residents and staff in response to outbreaks, and finally non-outbreak testing of asymptomatic HCP. The addition of non-outbreak testing may provide a modest benefit, although it requires more testing resources to prevent each COVID-19 case. The testing resources required (per case prevented) did decrease as the likelihood of SARS-CoV-2 introductions (i.e., level of community transmission) increased. The impact is still only modest even when SARS-CoV-2 introductions are more frequent than observed in the United States in July-September 2020. The reason that non-outbreak testing only has a modest benefit is that the benefit of non-outbreak testing relies on the ability to identify infected persons at key times in the course of their illness: the pre-symptomatic infectious period and, for asymptomatic individuals, the period of peak infectiousness. Both of these time periods are narrow (e.g., pre-symptomatic period of 2-3 days) and easily missed without very frequent (e.g., every-three-days or more often) testing [10,14]. Furthermore, implementing high-quality outbreak response leads to smaller outbreaks, which then reduces the added benefit of non-outbreak testing because the outbreaks prevented through non-outbreak testing are smaller in size.

The results described in this paper also support current recommendations that performing outbreak testing every three days is the preferred frequency when initiating an outbreak response, if this does not detract from other important activities such as infection control procedures [18]. In general, testing strategies with better turnaround time and higher frequency were estimated to be more effective if other conditions (e.g., infection control practices) remained constant.

The federal government is providing nursing homes with point-of-care antigen testing platforms [19]. The analysis conducted in this manuscript suggests that the advantage of faster turnaround time from such platforms may outweigh the disadvantage of potential lower sensitivity. Practical considerations may still limit how frequently these tests can be performed through serial testing strategies. An exploratory analysis suggested that a hypothetical strategy using daily point-of-care testing with lower (e.g., 50%) sensitivity could be as effective as, or even more effective than, relying on more sensitive tests used less frequently. Daily testing is likely not currently feasible in all nursing homes and would need widespread availability of inexpensive, simple testing options that could be run on easy-to-collect specimen types. In addition, making a test with 50% sensitivity available (that might facilitate daily testing) would require changes to the current regulatory framework [20–24].

These findings are subject to at least the following limitations. First, the analysis used mathematical modeling rather than directly studying an implementation. However, the model parameters have been reviewed as suitable to use for pandemic planning scenarios and are based on observational data about SARS-CoV-2 transmission [10]. The patterns seen and comparisons made from results, therefore, are likely reasonable even if the absolute estimates themselves may lack precision. Second, outbreaks in facilities have been assumed to be independent. Since some HCP work in more than one facility [4], the potential effectiveness of both testing strategies has been underestimated on a population level. Third, other unintended consequences of asymptomatic testing were not evaluated, such as identifying false positives during non-outbreak testing with population prevalence of COVID-19. Even if using a test with 99.4% specificity, most facilities without COVID-19 might expect at least one false positive result after testing all its staff. Fourth, there is uncertainty around the parameters used for the model. For example, the actual infectivity profile of SARS-CoV-2 infection is not known with certainty, although our main conclusions appear to be robust when evaluating several published infectivity profiles. As another example, the proportion of patients with asymptomatic infections has varied in the reported literature [25,26]. Fifth, we have not accounted for how the accumulation of persons with immunity over the course of time might affect strategies. Finally, if infection control practices, in general, are not optimal, then the benefits of testing strategies will be less than we have estimated. For example, we assume that outbreak testing occurs immediately after identification of a recognized case, while in practice delays could occur.

In summary, testing asymptomatic persons in response to an outbreak is an effective and efficient strategy to supplement optimized infection control practices to limit SARS-CoV-2 transmissions in nursing homes. The benefit of outbreak testing strategies may increase with more frequent outbreak testing with rapid turnaround times, but most importantly require careful effort to ensure that infection control measures are effectively implemented.

## Supporting information

Supplemental Methods and Results

## Data Availability

Data provided in supplemental materials or publicly available through links in the manuscript.

https://github.com/cdcepi/Nursing-home-SARS-CoV-2-testing-model/

## Acknowledgements

The authors would like to thank Christopher Czaja, MD, DrPH (Colorado Department of Public Health and Environment) for discussions about nursing home outbreak response in Colorado early in the COVID-19 pandemic.

## Conflicts of interest

None

## Funding

This work was funded by the Centers for Disease Control and Prevention.

## Disclaimer

The findings and conclusions in this report are those of the authors and do not necessarily represent the views and the official position of the Centers for Disease Control and Prevention.

## Notes

### Competing Interest Statement

The authors have declared no competing interest.

### Funding Statement

No external funding was received. All work was conducted as part of government duties.

### Author Declarations

This activity was reviewed by CDC and was conducted consistent with applicable federal law and CDC policy (see e.g., 45 C.F.R. part 46, 21 C.F.R. part 56; 42 U.S.C. 241(d); 5 U.S.C 552a; 44 U.S.C. 351 et seq.).

### Summary of Updates

New Table 1 added. Text revised at request of peer review. CDC epi GitHub link added.

