## Supplemental Methods and Results for "Modeling effectiveness of testing strategies to prevent COVID-19 in nursing homes —United States, 2020"

Baseline parameters:

For the analysis reported in the main text, we have used the updated infectivity profile estimated by He et al. (1), in which the infectiousness varies with time. At *x* days from symptom onset, infectiousness is described by a Gamma density function that is evaluated at time $x + t_0$ with shape *a* and scale *s,* where $\left(a, \frac{1}{s}\right) = (20.516508, 1.592124)$. We have also assumed CDC "best estimates" that 50% of transmissions occur before symptom onset, 60% of infections are symptomatic, a generation time of 6.5 days, and ratio of infectivity for asymptomatic:symptomatic infections is 0.75 during the course of illness from the September 10, 2020 update(2). The parameter $t_0$ was modeled using the Gamma quantile function with shape and scale parameters as above to be consistent with CDC modeling assumptions that 50% of transmissions from symptomatic cases occur during the presymptomatic phase, yielding $t_0 = 12.272481$.

Modeling effectiveness of preventing transmission

Let the infectiousness of an infected individual be $I$(t) (normalized to unity, so that $\int_0^\infty I(t)dt = 1$, where *t* is the time since exposure), and the fraction of infected individuals who have already shown symptoms at *t* be σ*(t)*. Then, the proportion of transmissions eliminated on isolation of symptomatic individuals for a period $t_I$ from symptom onset is $P_s = \int_0^\infty \sigma(t)[I(t) - I(t + t_I)]\, dt$. (If the time from beginning of infectiousness to onset of symptoms is believed to be fixed at $t_s$, the previous expression reduces to $P_s = \int_{t_s}^{t_s+t_I} \sigma(t) I(t) dt$. This is the version used in this analysis.) If an asymptomatic or presymptomatic infected individual is tested at time $t_0$ after infection [test sensitivity $s(t_0)$], and the test result is available after a reporting lag $t_I$ at $t_1 = t_0 + t_I$, the additional proportion of transmissions eliminated by isolation on return of a positive test result is

$$\rho_a(t_0, t_I) = s(t_0) \int_{t_1}^{t_1+t_I} [1 - \sigma(t)] I(t) dt \ .$$

If testing is done at regular intervals *T*, then the mean proportion of asymptomatic transmissions eliminated is

$$P_a(T, t_I) = \frac{1}{T} \int_0^T \{\rho_a(t + T, t_I) + [1 - s(t)]\rho_a(t + T, t_I) + [1 - s(t)][1 - s(t + T)]\rho_a(t + 2T, t_I) + \cdots\}\, dt.$$

The second term in the integrand accounts for those who tested false negative once, followed by a true positive; the third term accounts for those with two false negatives, etc. For convenient numerical evaluation, note that the integrand, $\pi(t, T, t_I)$ satisfies the recursive relation $\pi(t, T, t_I) = \rho_a(t, t_I) + [1 - s(t)]\pi(t + T, T, t_I)$.

For COVID-19 parameters used here, setting $\pi(t, T, t_I) = 0$ for $t > 20$ days was found to yield sufficient precision.

DRAFT

In these analyses $\sigma(t)$ after symptoms have occurred is considered constant and referred to as $f_{symp}$ below.

Modeling of outbreak testing

Publicly available data from the Colorado Department of Public Health and Environment (CDPHE) for outbreaks in congregate settings was obtained and limited to those occurring in nursing homes (3). Outbreaks ending prior to May 6, 2020 (approximately when local guidelines for testing began to be implemented, Chris Czaja, personal communication) were used as a representation of the distribution of outbreak size in nursing homes during our baseline period when testing and isolation only occurred based on symptoms. Other infection control precautions currently recommended (e.g., universal use of facemasks by HCP) were already recommended by CDC at the time. The data from CDPHE were linked to Centers for Medicare & Medicaid Nursing Home Compare (4) to find the number of beds at the facility. Total outbreak size was defined as both confirmed and suspected COVID-19 cases in residents or HCP. The final outbreak size was not correlated with the facility bed size, which suggested that models in which the baseline outbreak size was independent of facility size would be more appropriate than those based on a fixed attack rate at the facility.

A Reed-Frost model was selected to model an outbreak in this setting. Using the estimated number of people at facilities with outbreaks in Colorado, for each facility the $R_0$ value for which the mean outbreak size equaled the observed size was calculated. The unmitigated reproductive number $R_{0,unmit}$ for a typical outbreak unmitigated by testing strategies was estimated by taking the mean of the various estimated values of $R_{0,}$, which was 1.388867. As noted previously at baseline the reduction in transmission $\mu$ calculated above from only isolating/testing symptomatic persons is $trans_{symp,old}$ and after implementing a testing strategy is $trans_{symp,new}$. The new reproductive number when mitigated by testing: $R_{0,mit} = R_{0,unmit}\frac{1-trans\_asymp-trans\_symp\_new}{1-trans\_symp\_old}$. When evaluating testing strategies, outbreaks were considered to occur unmitigated by testing strategies for the first generation and then mitigated afterwards.

Modifications of additional parameters

*Change in effectiveness of infection control precautions*

The models above assume that once a person has been identified (through symptoms or testing), then isolation precautions are immediately instituted and the person no longer transmits SARS-CoV-2. The effect of incomplete isolation of persons was incorporated as follows. In the context of ideal infection control practices, the proportion of transmission prevented by detection and isolation of symptomatic patients is $trans_{symp}$. Instead of complete cessation of transmission through isolation only a

DRAFT

proportion of transmission of identified patients is prevented represented by a factor $isol_{eff}$, so with less effective isolation of patients the proportion of transmission prevented is now $trans_{symp}' = isol_{eff} \times trans_{symp}$.

Now as before we will have a new reproductive number from only symptom screening which is

$R_{0,unmit}' = R_0 \times \frac{1-trans_{symp}'}{1-trans_{symp}}$ where $R_0$ is the original reproductive number with ideal isolation precautions. As before the new $R_{0,mit}' = R_{0,unmit}' \times \frac{1-trans_{asymp}'-trans_{symp}'}{1-trans_{symp}}$ where $trans_{asymp}' = isol_{eff} \times trans_{asymp}$, to determine the size of outbreaks N' after conducting outbreak testing.

Because HCP detected by non-outbreak testing are assumed to be literally removed from the healthcare facility after identification through this strategy, no change to the results of the percentage of outbreaks prevented through non-outbreak testing was made based on changes to effectiveness of infection control precautions.

<u>Estimating effect of testing strategies</u>

To determine the percentage of infections prevented by incorporating a testing strategy for a facility not currently in the midst of an outbreak response, for outbreak testing, the proportional decrease $\delta_1$ in expected outbreak size for an unmitigated versus mitigated outbreak was calculated. When evaluating the testing strategy of combining outbreak and non-outbreak testing, the proportionate decrease $\delta_2$ in transmission for non-outbreak testing by itself was first calculated, which in our framework equals the proportional drop in number of outbreaks and hence the number of infections. Because outbreak testing leads to those outbreaks occurring to be smaller, the proportionate decrease in infections when using both outbreak testing and non-outbreak testing is

$$D = 1 - (1 - \delta_1)\,(1 - \delta_2)$$

To calculate the number of tests used per infection prevented, for non-outbreak testing the probability *h* that a facility would experience a new outbreak was varied, so that a facility with *N* persons (residents+HCP) and *M* HCP not experiencing an outbreak and conducting non-outbreak testing every *d* days would perform $M \times 7/d$ tests.

To determine a value for *h* to use for most calculations, data from CMS certified nursing homes reporting to the National Healthcare Safety Network were analyzed. For each week from July – September 2020, among nursing homes reporting no confirmed COVID-19 cases among residents or staff in the previous four weeks, the percentage of facilities reporting a confirmed case ranged from 9.1% – 14.1% (median: 12.1%). Based on these findings, *h* was set at 10% unless otherwise specified.

For outbreak testing, the mean number of generations from the Reed-Frost model was calculated and converted to a number of weeks duration of the outbreak using 6.5 days as the typical generation time. Serial testing at the specified frequency occurred through the duration of the outbreak and then for an additional two weeks facility-wide testing to confirm no new infections occurred.

DRAFT

Then the expected total number of infections prevented per facility is $40 \times D \times h$

Expected number of tests per facility is the number of tests needed for outbreak testing $N \times (L + 2) \times 7/d$ times the likelihood of an outbreak occurring $h \times (1 - \delta_2)$, plus the number of non-outbreak tests needed:

$$N\ [h(L + 2)\ (1 - \delta_2)\ \times\ 7/d]\ +\ M \times 7/d$$

Where *L* is the expected number of weeks for a mitigated outbreak.

For analyses in which the effectiveness of isolating patients has been reduced, the results of the expected size of outbreaks from less effective isolation in conjunction with outbreak and non-outbreak testing are compared to the size of outbreaks from effective isolation and only symptom screening to determine the percent reduction in number of infections.

For analyses in which the ascertainment of symptoms was reduced, we compared the number of infections occurring with symptom screening alone, with outbreak testing alone, and with the combination of both outbreak and non-outbreak testing; reduction of ascertainment of symptoms was applied to all three scenarios.

<u>Sensitivity analyses</u>

As a sensitivity analysis to ensure that results are not tied to the specific infectivity profile chosen from He et al, the analyses of percent of cases prevented and number of tests per case prevented at a 10% probability of SARS-CoV-2 introduction were conducted. We employed four additional infectivity profiles based on available modeling studies: an alternative infectivity profile using the Gamma distribution based on parameters from a combination of multiple studies [4-8] as described in [9], and adaptation of the infectivity profile published by Goyal et al [10], and adaptation of the infectivity profile published by Clifford et al [11]. A summary of the infectivity profiles used is described in supplemental table S1 and displayed on the supplemental Figure, and results from these sensitivity analyses can be found supplemental tables S2-S4.

<u>Additional limitations with the analysis are as follows:</u>

We do not have data to confidently determine the correct distribution of the HCP:resident ratio. In addition, we have estimated the impact of different testing strategies for a “typical” facility although the numbers of residents will vary in reality.

In terms of the modeling, many additional refinements are possible such as modeling HCP and residents as separate compartments or individual-level modeling.

For calculating the expected number of tests per facility per infection prevented, the calculation has been simplified by treating facilities with introductions leading to outbreaks as a separate group from those without, whereas in reality multiple processes may occur at once, e.g., new introductions could occur while facilities are finishing a previous outbreak. In addition, there is a difference in follow-up time when calculating testing needs for a facility having a SARS-CoV-2 introduction leading to an

DRAFT

outbreak which has not been accounted for directly. These areas could be refined in future explorations of SARS-CoV-2 testing in congregate settings.

We have assumed a constant generation time. If infection control practices change over time the actual generation time in practice may not be constant.

We have also only modeled the situation using a single $R_0$ value. If the true $R_0$ within a facility is much larger then we will have underestimated the benefit of non-outbreak testing. In addition, we did not model situations for smaller $R_0$ values that hypothetically might occur (for example in the presence of an effective and widely-used vaccine in nursing homes), as this was outside the scope of this paper.

Finally, we do not know for certain how much the infection control practices and symptom screening in nursing homes in Colorado may or may not have conformed to ideal practices or whether these are truly representative of a typical facility. Experience with nursing homes has suggested that symptom ascertainment may be quite complete (CDC, unpublished data). If the observed outbreak sizes we use do not conform to optimal infection control practices then we may be even more severely underestimating the potential benefit of excellent infection control in nursing homes to prevent SARS-CoV-2 infections in that setting. We also may overestimate some of the benefits of adding testing strategies if baseline practices for isolation are not good.

DRAFT

**Supplemental Table S1: SARS-CoV-2 infectivity profiles used**

| Profile reference | Description | Location of results in paper |
|---|---|---|
| He et al [1] | Updated parameters for gamma density function, (shape 20.5165082 and 1/scale 1.5921240) | Table (main text) |
| Combination of multiple studies [4-8] as described in Johansson et al [9] | Gamma density function with ten-day infectious period and peak infectiousness on day 5 (shape 7.2, scale 0.8) | Table S2 |
| Goyal et al [10] | Most people are infectious from days 3 to 7, with tapering afterwards | Table S3 |
| Clifford et al [11] | Based on estimated latent periods (the delay between infection and becoming infectious) [1] and infectious periods [12] | Table S4 |

The Goyal et al. and Clifford et al. infectiousness models were fitted by first simulating daily individual-level infectiousness profiles from the original code and then fitting density functions to the simulation data as described in [9]

DRAFT

**Supplemental Table S2**

Sensitivity analysis using infectivity profile from Johansson et al [9] modeled after the combination of parameters from various published studies [4-8] using gamma distribution (Table S1). Percent SARS-CoV-2 infections prevented and number of tests needed per case prevented when serially testing asymptomatic nursing home residents and staff in response to outbreaks ("outbreak testing"), combining this testing strategy with serial testing of asymptomatic staff in the absence of known cases ("outbreak+non-outbreak testing"), and conducting both outbreak and non-outbreak testing with a concomitant 10% decrease in the effectiveness of halting SARS-CoV-2 transmission through isolating identified cases. Cases occurring using these strategies are compared to the number occurring with use of only symptom-based testing and isolation[a].

| | | | Outbreak testing[b] | | Outbreak + non-outbreak testing[b,c] | | Outbreak + non-outbreak testing[b,c], 10% decrease in effectiveness of isolation | |
|---|---|---|---|---|---|---|---|---|
| **Sensitivity[d]** | **Turnaround time** | **Testing frequency** | **% cases prevented[e]** | **Tests/case prevented[e]** | **% cases prevented[e]** | **Tests/case prevented[e]** | **% cases prevented[e]** | **Tests/case prevented[e]** |
| Baseline | 48 hours | Weekly | 30.6% | 136 | 36.9% | 173 | 17.8% | 377 |
| Baseline | 24 hours | Weekly | 48.7% | 77 | 56.3% | 103 | 39.3% | 157 |
| Baseline | 48 hours | Every 3 days | 58.4% | 139 | 66.2% | 191 | 51.1% | 265 |
| Baseline | 24 hours | Every 3 days | 78.5% | 82 | 85.2% | 123 | 77.1% | 145 |
| 85% | POC | Weekly | 60.1% | 57 | 67.9% | 79 | 53.3% | 108 |
| 85% | POC | Every 3 days | 86.2% | 64 | 91.9% | 101 | 87.8% | 111 |
| 50%[f] | POC | Weekly | 38.6% | 104 | 45.7% | 134 | 27.3% | 238 |
| 50%[f] | POC | Every 3 days | 70.8% | 101 | 78.2% | 145 | 67.0% | 183 |
| 50%[f] | POC | Daily | 90.7% | 158 | 95.7% | 259 | 94.1% | 272 |

POC: point-of-care (i.e., results available immediately)

[a] In this situation a mean of 50 cases/outbreak occur.

DRAFT

[b] Outbreak testing: serial testing of asymptomatic persons in response to an outbreak

[c] Non-outbreak testing: serial testing of asymptomatic healthcare personnel in the absence of known cases

[d] Sensitivities of 85% and 50% are relative to the sensitivity of the baseline test, which is modeled to vary during the course of infectiousness and to replicate sensitivity of existing reverse transcriptase polymerase chain reaction (RT-PCR) tests for SARS-CoV-2.

[e] The fraction of cases prevented compared to what is expected to occur when performing neither outbreak testing nor non-outbreak testing but still maintaining effectiveness of isolating patients.

[f] No point-of-care test with this sensitivity is currently available for use in the United States; this testing platform represents a hypothetical situation in that respect or may be a reflection of tests that may have low sensitivity for detecting asymptomatic persons with low viral load.

DRAFT

**Supplemental Table S3**

Sensitivity analysis based on infectivity profile described in Goyal et al (Table S1) [10]. Percent SARS-CoV-2 infections prevented and number of tests needed per case prevented when serially testing asymptomatic nursing home residents and staff in response to outbreaks (“outbreak testing”), combining this testing strategy with serial testing of asymptomatic staff in the absence of known cases (“outbreak+non-outbreak testing”), and conducting both outbreak and non-outbreak testing with a concomitant 10% decrease in the effectiveness of halting SARS-CoV-2 transmission through isolating identified cases. Cases occurring using these strategies are compared to the number occurring with use of only symptom-based testing and isolation[a].

| | | | **Outbreak testing[b]** | | **Outbreak + non-outbreak testing[b,c]** | | **Outbreak + non-outbreak testing[b,c], 10% decrease in effectiveness of isolation** | |
|---|---|---|---|---|---|---|---|---|
| **Sensitivity[d]** | **Turnaround time** | **Testing frequency** | **% cases prevented[e]** | **Tests/case prevented[e]** | **% cases prevented[e]** | **Tests/case prevented[e]** | **% cases prevented[e]** | **Tests/case prevented[e]** |
| Baseline | 48 hours | Weekly | 43.9% | 88 | 51.3% | 116 | 33.4% | 190 |
| Baseline | 24 hours | Weekly | 63.4% | 53 | 71.1% | 73 | 57.3% | 98 |
| Baseline | 48 hours | Every 3 days | 73.4% | 94 | 80.6% | 138 | 70.2% | 170 |
| Baseline | 24 hours | Every 3 days | 86.4% | 63 | 92.0% | 100 | 88.0% | 110 |
| 85% | POC | Weekly | 72.8% | 41 | 80.0% | 60 | 69.5% | 74 |
| 85% | POC | Every 3 days | 90.4% | 54 | 95.4% | 88 | 93.6% | 92 |
| 50%[f] | POC | Weekly | 50.5% | 74 | 58.2% | 98 | 41.3% | 148 |
| 50%[f] | POC | Every 3 days | 80.5% | 77 | 86.9% | 117 | 79.8% | 136 |
| 50%[f] | POC | Daily | 92.6% | 143 | 97.3% | 235 | 96.5% | 242 |

POC: point-of-care (i.e., results available immediately)

[a] In this situation a mean of 50 cases/outbreak occur.

[b] Outbreak testing: serial testing of asymptomatic persons in response to an outbreak

DRAFT

[c] Non-outbreak testing: serial testing of asymptomatic healthcare personnel in the absence of known cases

[d] Sensitivities of 85% and 50% are relative to the sensitivity of the baseline test, which is modeled to vary during the course of infectiousness and to replicate sensitivity of existing reverse transcriptase polymerase chain reaction (RT-PCR) tests for SARS-CoV-2.

[e] The fraction of cases prevented compared to what is expected to occur when performing neither outbreak testing nor non-outbreak testing but still maintaining effectiveness of isolating patients.

[f] No point-of-care test with this sensitivity is currently available for use in the United States; this testing platform represents a hypothetical situation in that respect or may be a reflection of tests that may have low sensitivity for detecting asymptomatic persons with low viral load.

DRAFT

**Supplemental Table S4**

Sensitivity analysis based on infectivity profile described in Clifford et al (Table S1) [11]. Percent SARS-CoV-2 infections prevented and number of tests needed per case prevented when serially testing asymptomatic nursing home residents and staff in response to outbreaks (“outbreak testing”), combining this testing strategy with serial testing of asymptomatic staff in the absence of known cases (“outbreak+non-outbreak testing”), and conducting both outbreak and non-outbreak testing with a concomitant 10% decrease in the effectiveness of halting SARS-CoV-2 transmission through isolating identified cases. Cases occurring using these strategies are compared to the number occurring with use of only symptom-based testing and isolation[a].

| | | | Outbreak testing[b] | | Outbreak + non-outbreak testing[b,c] | | Outbreak + non-outbreak testing[b,c], 10% decrease in effectiveness of isolation | |
|---|---|---|---|---|---|---|---|---|
| **Sensitivity[d]** | **Turnaround time** | **Testing frequency** | **% cases prevented[e]** | **Tests/case prevented[e]** | **% cases prevented[e]** | **Tests/case prevented[e]** | **% cases prevented[e]** | **Tests/case prevented[e]** |
| Baseline | 48 hours | Weekly | 74.0% | 40 | 81.1% | 58 | 70.8% | 72 |
| Baseline | 24 hours | Weekly | 81.6% | 32 | 87.9% | 49 | 81.2% | 56 |
| Baseline | 48 hours | Every 3 days | 88.3% | 59 | 93.7% | 94 | 90.7% | 102 |
| Baseline | 24 hours | Every 3 days | 91.4% | 51 | 96.3% | 84 | 95.0% | 87 |
| 85% | POC | Weekly | 83.2% | 30 | 89.3% | 47 | 83.5% | 53 |
| 85% | POC | Every 3 days | 92.3% | 49 | 97.0% | 80 | 96.1% | 83 |
| 50%[f] | POC | Weekly | 64.8% | 51 | 72.5% | 71 | 58.8% | 94 |
| 50%[f] | POC | Every 3 days | 86.8% | 63 | 92.3% | 99 | 88.5% | 109 |
| 50%[f] | POC | Daily | 93.6% | 134 | 98.2% | 219 | 97.8% | 224 |

POC: point-of-care (i.e., results available immediately)

[a] In this situation a mean of 50 cases/outbreak occur.

[b] Outbreak testing: serial testing of asymptomatic persons in response to an outbreak

DRAFT

[c] Non-outbreak testing: serial testing of asymptomatic healthcare personnel in the absence of known cases

[d] Sensitivities of 85% and 50% are relative to the sensitivity of the baseline test, which is modeled to vary during the course of infectiousness and to replicate sensitivity of existing reverse transcriptase polymerase chain reaction (RT-PCR) tests for SARS-CoV-2.

[e] The fraction of cases prevented compared to what is expected to occur when performing neither outbreak testing nor non-outbreak testing but still maintaining effectiveness of isolating patients.

[f] No point-of-care test with this sensitivity is currently available for use in the United States; this testing platform represents a hypothetical situation in that respect or may be a reflection of tests that may have low sensitivity for detecting asymptomatic persons with low viral load.

DRAFT

**Supplemental Figure:** SARS-CoV-2 infectiousness over time for infectivity profiles used (described in table S1).

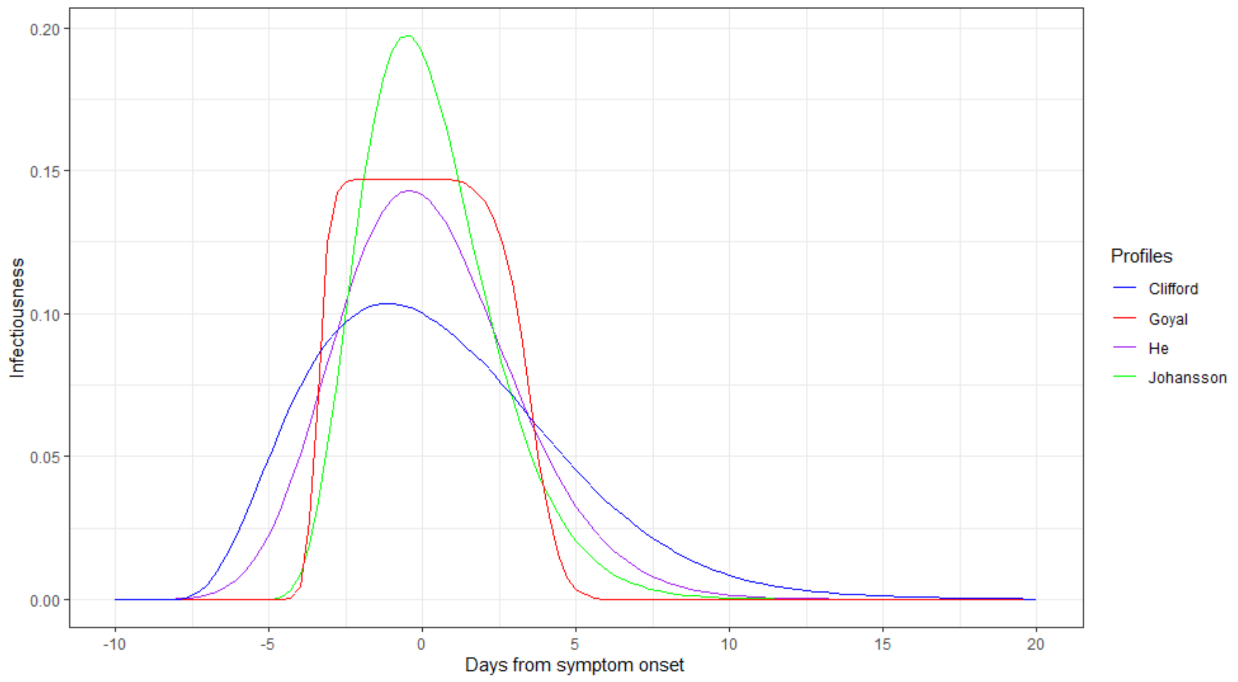


DRAFT
